# Novel blood test for early biomarkers of preeclampsia and Alzheimer’s disease

**DOI:** 10.1101/2021.03.19.21253961

**Authors:** Shibin Cheng, Sayani Banerjee, Lori A. Daiello, Akitoshi Nakashima, Sukanta Jash, Zheping Huang, Jonathan D. Drake, Jan Ernerudh, Goran Berg, James Padbury, Shigeru Saito, Brian R. Ott, Surendra Sharma

**Author notes:** Address correspondence to Surendra Sharma, MD, PhD, Department of Pediatrics, Women and Infants Hospital, 101 Dudley Street, Providence, Rhode Island 02905, USA. S.C. and S.B. contributed equally to this work.

## Abstract

Non-invasive and sensitive blood test has long been a goal for early stage disease diagnosis and treatment for Alzheimer’s disease (AD) and other proteinopathy diseases. However, a blood test based on a mechanistic link to pathologic protein aggregate complexes has not been yet elucidated. We previously reported that preeclampsia (PE), a severe pregnancy complication, is another proteinopathy disorder with impaired autophagy. We hypothesized that induced autophagy deficiency would promote accumulation of pathologic protein aggregates. Here, we describe a novel, sensitive assay that detects serum protein aggregates from patients with PE as well as AD in both dementia and prodromal mild cognitive impairment (MCI) stages. The assay employs exposure of genetically engineered, autophagy-deficient human trophoblasts (ADTs) to serum from patients. The aggregated protein complexes and their individual components, including transthyretin, amyloid β-42, α-synuclein, and phosphorylated tau231, can be detected and quantified by co-staining with ProteoStat, a rotor dye with affinity to aggregated proteins, and respective antibodies. Autophagy-proficient human trophoblasts failed to accumulate serum protein aggregates under similar culture conditions. Detection of protein aggregates in ADTs was not dependent on transcriptional upregulation of these biomarkers. The ROC curve analysis validated the robustness of the assay for its specificity and sensitivity. In conclusion, we have developed a novel noninvasive diagnostic and predictive assay for AD, MCI and PE.

## Introduction

Toxic extracellular and intracellular deposition of misfolded protein aggregates in the brain is a hallmark feature of proteinopathy in many neurodegenerative diseases such as Alzheimer’s (AD) ^1–4^. Additionally, we and others have recently demonstrated that preeclampsia (PE), a severe pregnancy complication, mechanistically exists on the spectrum of proteinopathies^5–11^. PE is diagnosed by presentation with *de novo* onset of hypertension and proteinuria at or after 20 weeks of gestation^12–15^. PE represents a unique case of proteinopathy in younger population and can also lead to subsequent chronic conditions later in life^16–18^, including mild cognitive impairment and dementia in mothers^18–21^ and their offspring^22^.

Both AD and PE are devastating disorders for which there are currently no rapid, non-invasive methods available for detection of protein aggregates in the prodromal phase of AD or at early stage of pregnancy. In the case of AD, pathophysiological protein aggregation is thought to begin in a preclinical phase at least a decade prior to the onset of even subtle clinical symptoms, and then gradually progress through prodromal mild cognitive impairment (MCI) and clinical stages of AD dementia^23^. The definite diagnosis of AD is defined by seeded growth and histopathological evidence of extracellular amyloid β (Aβ) plaques and intracellular neurofibrillary tangles involving diverse hyperphosphorylated tau isoforms in the post-mortem brain^24, 25^. The probabilistic diagnosis of AD has been historically defined by elevated levels of Aβ in cerebrospinal fluid (CSF) in individuals with AD or MCI^26–28^. Additionally, although initially characterized as pathological changes in Parkinson’s disease and Lewy body dementia, recent observations suggest that α-synuclein (α-syn) is deposited concomitantly with the AD pathology in a high number of cases^29–31^. Evidence also exists for reduced levels of choroid plexus-derived transthyretin (TTR), transporter of thyroxine and retinol, in CSF and plasma of AD patients^32–35^. Thus, a blood test should be able to score for protein aggregate complexes containing all these biomarker components.

The currently available and validated tests mainly depend on detection of Aβ or phosphorylated tau in CSF or brain using mass spectrometry or positron emission tomography (PET) imaging^24, 36–43^. However, the invasive nature of CSF-based measurements and the cost of PET imaging make these tests less attractive and time consuming. Efforts have recently been reported on the development of plasma-based tests that involve Aβ immunoprecipitation and mass spectrometry analysis or high plasma levels of P-tau181, P-tau217 or P-tau231^39–42^. Although of great significance, these plasma tests depend on detection of a monomeric or single protein, require significant amounts of blood, and may involve biochemical manipulation of plasma.

Here we leverage the concept that autophagy removes aggregated proteins and damaged organelles from cells. We describe a novel blood test that utilizes autophagy-deficient human trophoblasts as “targets” to engulf and accumulate protein aggregates. We have successfully detected total protein aggregates and their specific components Aβ, TTR, P-tau231, and α-syn in sera from patients with AD, MCI, and PE. Our blood-based assay can broadly and accurately detect serum aggregated protein biomarkers in proteinopathy diseases.

## Results

### Novel strategy and its validation for detecting protein aggregates

Since a number of proteins are likely to be involved in the pathological progression of proteinopathy diseases, we designed a strategy to detect multiple proteins and their aggregates. Our strategy is based on the observation that protein aggregates accumulate in human trophoblasts when autophagy is impaired by induction of endoplasmic reticulum stress^11^. Prior studies have shown that trophoblast cells have strong endocytic and phagocytic ability to internalize large molecules and complexes^46, 47^. We hypothesized that protein aggregate complexes would be readily endocytosed but not easily degraded or cleared by autophagy deficient trophoblasts (ADTs). To test this hypothesis, we established an ADT cell line using human first trimester extravillous trophoblasts by stably transfecting and expressing ATG4BC74A, an inactive mutant autophagy gene of ATG4B (see Methods for details) ^45^. The mutant ATG4B^C74A^ inhibits conversion of LC3-I to LC3-II and subsequently blocks autophagic flux (Fig. 1a). The engineered ADTs were confirmed to exhibit not only disrupted autophagic flux but also impaired lysosomal biogenesis in our prior studies^11^. We then examined whether ADTs can take up and accumulate protein aggregates. Aggregated transthyretin (TTR) has been detected in proteinopathy diseases, including preeclampsia^5^. An *in vitro* method to induce TTR aggregate formation is described in Methods. Using this *in vitro* approach, we first generated human TTR aggregates and verified the conversion of TTR into aggregates by a rapid optimal density (OD) analysis that detects altered fluorescence readings for dye-aggregate complexes (Supplementary Fig. 1). Based on analysis of the fluorescence signals, we could determine the *in vitro* aggregation index of TTR (Supplementary Fig. 1). For the cell-based assay, ADTs were incubated on ice with *in vitro* generated TTR aggregates to allow synchronization of entry. After 20 min-incubation, the cells were either immediately fixed (referred to as 0 h time point) or incubated at 37 °C for 24 h. As controls, the cells were incubated with native (non-aggregated) TTR in parallel. The fixed cells were then stained with ProteoStat, a rotor dye with affinity to aggregated protein structures. In ADTs treated with aggregated TTR at 0 h, ProteoStat-positive clump-like aggregates were distributed on the surface and in extracellular areas of the cells. After 24 h incubation, robust ProteoStat fluorescent signal for TTR aggregates was readily detected in almost all cells in the well. However, no significant ProteoStat signal was observed in ADTs incubated with native TTR (Fig. 1b). These observations validate the concept that aggregated protein complexes will accumulate in ADTs and can be easily detected by using ProteoStat. This novel method provides a basis for detecting protein aggregates in serum and in fluids such as serum and urine.

**Figure 1.**
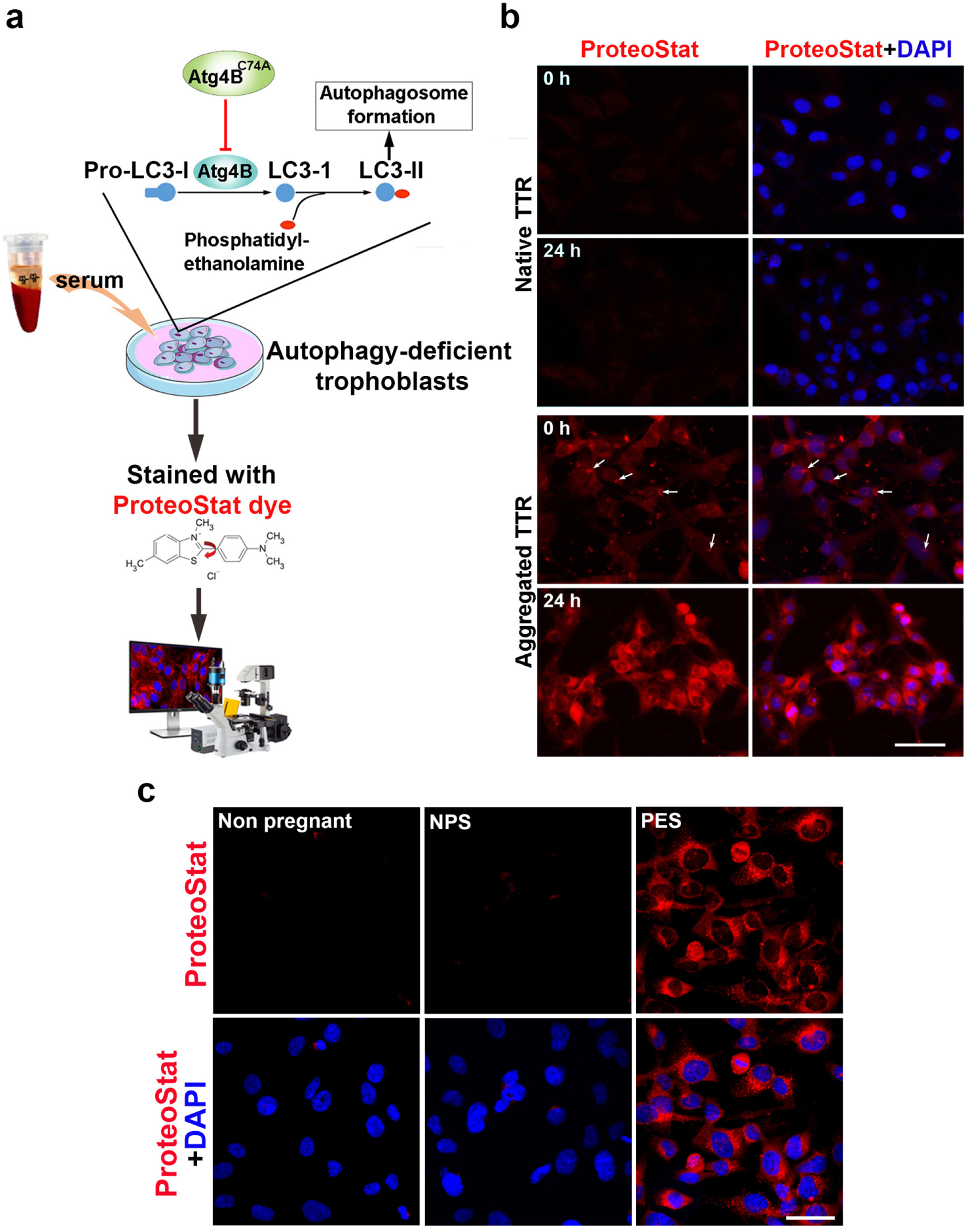
Validation of the protein aggregation assay. (A) Schematic diagram of the protocol. Autophagy-deficient trophoblast cells (ADTs) were incubated for various time periods with aggregate-containing sera, fixed and then stained with ProteoStat dye alone or in combination with immunostaining with specific protein antibodies. Red ProteoStat fluorescent signal was imaged with laser confocal microscope and then quantified. (B) Representative images for the internalization and accumulation of *in vitro* generated transthyretin (TTR) aggregates in ADTs. TTR aggregates were generated by incubation of recombinant human TTR in acetate buffer (see Materials and Methods). ADTs were treated with native TTR or aggregated TTR, fixed at 0 h or overnight after 20-min incubation on ice and then stained with ProteoStat dye (red). The nuclei were stained with DAPI (blue). Bar: 20 µm. (C) Incubation of sera from early onset preeclampsia (PES) not from non-pregnancy and normal pregnancy (NPS) results in accumulation of total protein aggregates in ADTs. Images are representatives of at least 3 independent experiments. The nuclei were stained with DAPI (blue). Bar: 50 µm.

### Serum-based detection of protein aggregates

In pilot experiments, we tested sera from early (e-PES) and late (l-PES) onset PE patients as well as gestational age-matched normal pregnancy control serum samples (Supplementary Table 1). As described in Materials and Methods, PE diagnosis followed the American College of Obstetricians and Gynecologists (ACOG) guidelines. We initially analyzed serum samples from non-pregnant women (n = 4), normal pregnancy (n = 4), and e-PE (n = 4). ADTs grown on cover slips were incubated with serum samples for 24 h. Cells were fixed and stained with ProteoStat (see Methods). Cytoplasmic accumulation of protein aggregates was confirmed by ProteoStat fluorescence signal as DAPI-stained nuclei were almost devoid of any fluorescence signal. Data shown in Fig. 1c strongly suggest that ADTs can be used to detect serum protein aggregates from PE patients.

To generalize the ADT-ProteoStat protein aggregate detection assay and to assess the kinetics of accumulation of protein aggregates, we incubated ADTs with FBS-free medium supplemented with sera from women with e-PE (n = 33), l-PE (n = 33), or respective gestational age-matched normal pregnancy controls (n = 39 for e-PE, n = 38 for l-PE). Cells were fixed at indicated time points and then stained with ProteoStat dye. The pixel intensity of the ProteoStat signal, as analyzed under a confocal microscope, was measured with ImageJ software and statistically compared among different groups at various time points. As shown in Fig. 2a, ProteoStat-positive protein aggregates started to appear in a few cells treated with e-PE serum (e-PES) even at 1 h. The total number of aggregate-containing ADTs and intensity of the ProteoStat signal increased in a time-dependent manner. At 24 h, almost all cells (> 85%) displayed robust ProteoStat signal. In contrast, only a very weak signal was observed in cells exposed to sera from gestational age-matched controls even at 24 h (Fig. 2a). With l-PE serum exposure, only a small number of cells showed fluorescent signal at 6 h, the intensity of which was lower than that with e-PES treatment at 6 h. However, at 24 h, about 60% of cells exhibited relatively stronger ProteoStat signal (Fig. 2b). Quantitative analysis revealed a significant increase in the pixel intensity of the ProteoStat fluorescence signal in ADTs incubated with either e-PES or l-PES vs. respective controls at 24 h (Fig. 2c, *p* < 0.01). The increased abundance of protein aggregates in e-PES and l-PES compared to control serum samples is reflected by the high AUC’s on ROC analysis of 1.0, *p* < 0.001 and CI of 0.949-1.000 (Fig. 2d). To better distinguish e-PE from l-PE, a careful screening of a larger set of samples at earlier kinetic time points may be needed.

**Figure 2.**
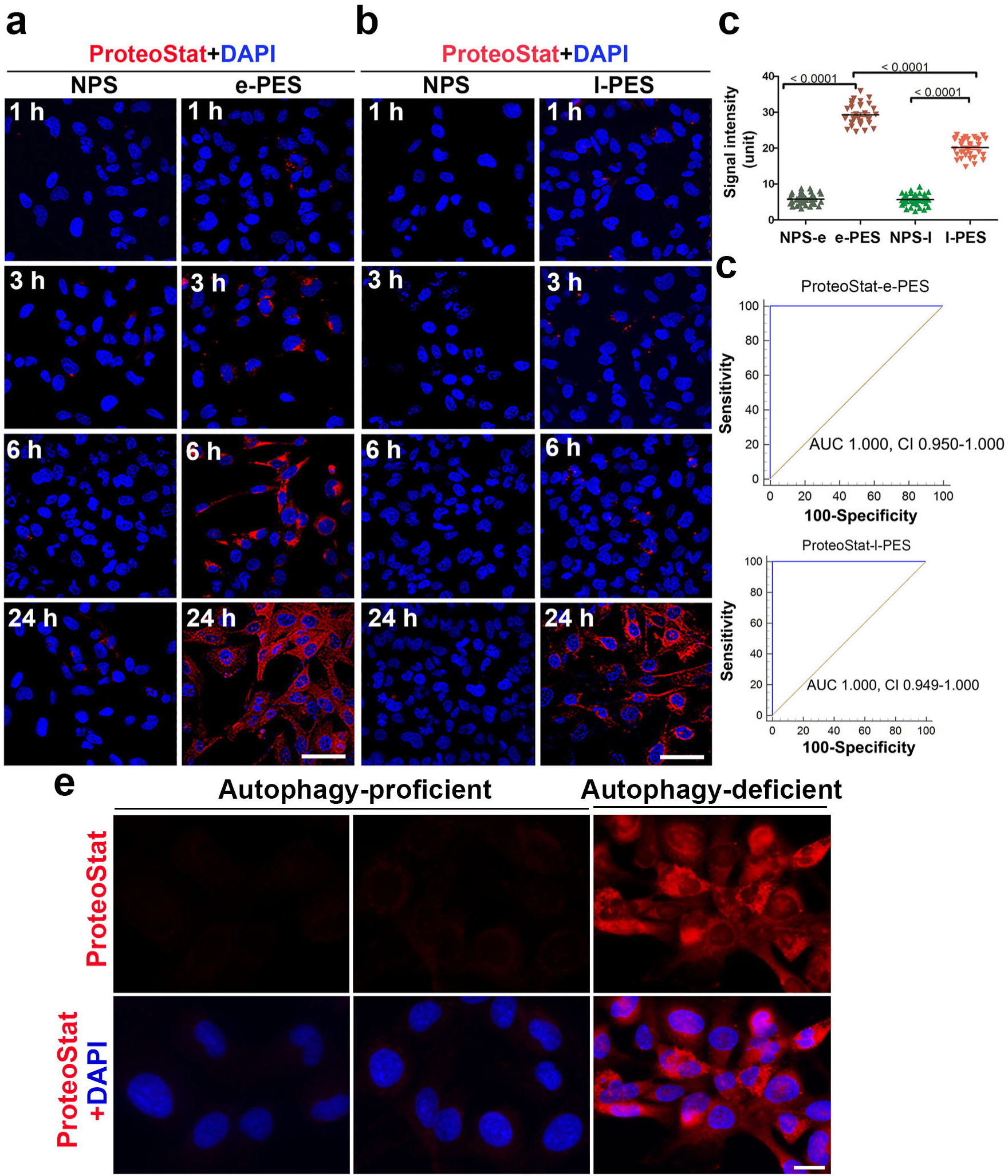
Kinetic accumulation of protein aggregates in ADTs treated with sera from women with e-PE, l-PE or respective controls. (A, B) Cells were grown in the media supplemented with 10% of early preeclampsia sera (e-PES) (n = 33)/control (n = 39) (A), or late preeclampsia sera (l-PES) (n = 33)/control (n = 38) (B), fixed at various time points and stained with ProteoStat dye (red). Shown are representative images. Bar: 50 µm. (C) Comparison of protein aggregates in sera from women with early-PE (e-PES, n = 33), late-PE (l-PES, n = 33) and respective control (NPS-e for e-PES, n = 39; NPS-l for l-PES, n = 38). (D) ROC curve analysis of the abundance of protein aggregates for prediction of e-PE (n = 33) and l-PE (n = 33). Data are presented as mean ± SEM and analyzed by one-way ANOVA with Bonferroni post hoc test. (E) No accumulation of protein aggregates was observed in autophagy-proficient human trophoblasts when incubated with sera from preeclampsia. The nuclei were stained with DAPI (blue). Bar: 20 µm.

To further demonstrate the significance of ADTs for the detection of protein aggregates, we compared autophagy-proficient trophoblasts (APTs) and ADTs under the identical conditions of incubation for 24 h. As depicted in Fig. 2e, APTs exhibited no or very weak ProteoStat fluorescence signal even after 24 h exposure to e-PES as compared to ADTs.

We next determined whether protein aggregates detected in serum-exposed ADTs are directly derived from PES. To address this, we first depleted protein aggregates from serum samples by filtering them through a nitrocellulose membrane with a pore size of 0.22 μm. ADTs were then incubated with filtered e-PES or unfiltered serum sample from the same patient(s). Filtration of PES through a nitrocellulose membrane capable of trapping large-sized protein structures resulted in nearly complete depletion of protein aggregates as depicted by a very poor ProteoStat signal in ADT cells (Supplementary Fig. 2), suggesting that accumulated protein aggregates detected in ADTs originated in sera from PE patients. In contrast, normal pregnancy serum (NPS) from controls showed no ProteoStat signal.

### Identification of protein components of the aggregate complex in sera from PE patients

To identify specific proteins in aggregate complexes in PES, we performed co-localization staining using specific antibodies in combination with the ProteoStat dye. Our previous studies have suggested the presence of TTR aggregates in the placenta from PE patients^5^. Studies by others have shown the presence of SERPINA1 in urine from severe PE patients^6^. We also explored whether PE and other proteinopathy diseases such as AD shared any protein aggregate markers. Based on our previous observations on TTR and other findings, we interrogated whether TTR, Aβ, α-synuclein (α-syn) and SERPINA1 are among the components of protein aggregates in PES-treated ADTs. Our results showed that a large amount of TTR and Aβ, as well as robust ProteoStat fluorescence, were detected in e-PES-treated cells. Importantly, both TTR and Aβ immunoreactive signals were co-localized with the ProteoStat signal, indicative of aggregated nature of TTR and Aβ (Fig. 3a). Similar results were obtained for l-PES (Supplementary Fig. 3). By comparison, little or no TTR and Aβ ProteoStat signals were seen in cells treated with gestational age-matched control sera. Importantly, no immunoreactive SERPINA1 or α-syn were observed in either e-PES/l-PES-treated or control serum-treated ADTs, although robust ProteoStat signal was present in e-PES/l-PES-exposed ADTs (data shown only for e-PES in Supplementary Fig. 4). The pixel intensity of TTR and Aβ immunoreactive signals was measured with ImageJ and statistically compared among the groups. The quantitative analysis showed higher levels of TTR and Aβ in e-PES compared to l-PES (Fig. 3b). Like the highly significant ROC curve AUC and CI values for ProteoStat analysis, these values demonstrated an AUC of 1.0 and a CI of 0.912-1.000 (Fig. 3c).

**Figure 3.**
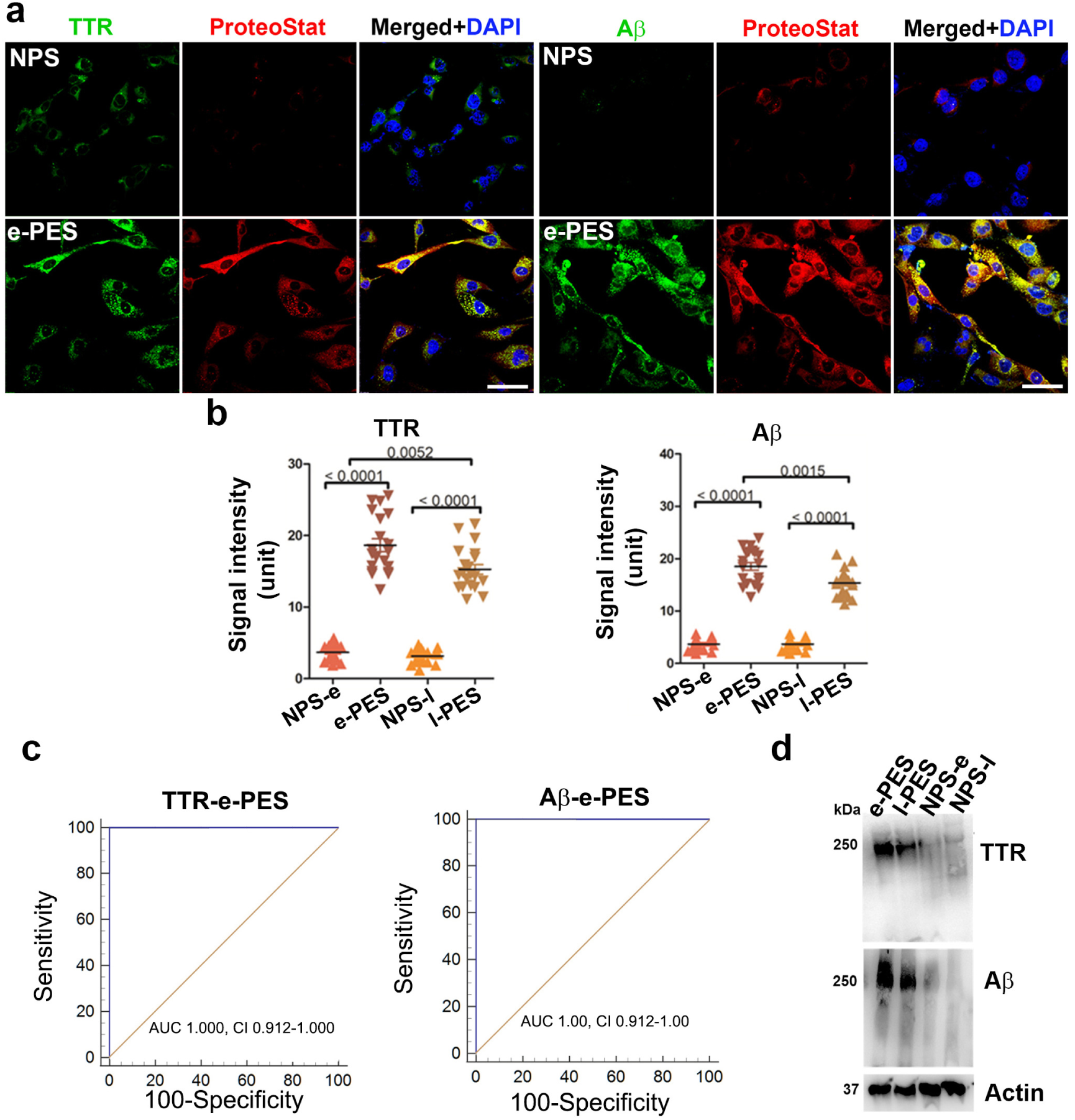
Identification of protein components of the aggregates in PE serum using co-localization staining with specific antibodies and ProteoStat dye and immunoblotting. ADTs were incubated with sera from e-PE (e-PES) or normal pregnancy controls and fixed at 24 h. (A) Fixed cells were serially stained with anti-TTR (green) or anti-Aβ antibodies (green) and ProteoStat dye (red). The nuclei were stained with DAPI (blue). Bar: 50 µm. (B) Quantitative analysis of signal intensity of TTR and Aβ in NPS- and e-PES/l-PES-treated cells. (C) ROC curve analysis of TTR and Aβ aggregates in sera from patients with e-PE and l-PE. Dual immunostaining for TTR and Aβ in combination with ProteoStat staining was performed in ADTs incubated with e-PE sera (e-PES), l-PE sera (l-PES) or respective control sera. The intensity of TTR and Aβ immunoreactive signals that were co-localized with ProteoStat fluorescence was measured and plotted for ROC curve. (D) Protein extracts were separated using western blotting under native conditions. The blots were then probed for TTR and Aβ. Actin was used as a loading control. Experiments were repeated at least 3 times.

To validate the findings described above, we utilized western blotting under native conditions to directly separate protein aggregates from extracts from the ADTs exposed to e-PES, l-PES or respective controls. As shown in Fig. 3d, a large amount of protein aggregates with high molecular weight was identified by anti-TTR and -Aβ antibodies in the ADTs treated with e-PES or l-PES but not from cells treated with corresponding control sera. These results are consistent with those obtained from the co-localization staining, indicating that TTR and Aβ, not α-syn and SERPINA1, are components of aggregated protein complexes in PE sera.

### Application of ADT-ProteoStat assay to sera from AD and MCI patients

To test the applicability of this ADT-ProteoStat assay for other well-known proteinopathy diseases, we investigated whether this assay can be employed to detect protein aggregates in sera from patients with AD and MCI. Participant characteristics describing baseline demographics, clinical assessments, imaging measures and fluid biomarker information are shown in Supplementary Table 2. Dementia severity was classified according to the Clinical Dementia Rating scale (CDR), a widely used and validated clinical scale that stratifies cognitive and functional impairment into group 0 (normal), group 0.5 (very mild cognitive impairment or questionable dementia), or groups 1-3 (mild to severe dementia)^48^. Biomarker support for AD pathology was available for 12/14 MCI cases. The two other cases lacked biomarker tests as part of their clinical diagnostic evaluations. These cases were highly likely to have AD pathology, however, as previous research has shown that people with CDR 0.5 or MCI almost always have AD pathology post-mortem^49, 50^. Biomarker support for AD pathology was available for 6/10 probable AD cases diagnosed according to National Institute on Aging-Alzheimer’s Association research criteria^51^. The four other cases lacked biomarker tests as part of their clinical diagnostic evaluations, but they also conformed to the National Institute on Aging-Alzheimer’s Association research criteria. Our overall design was to compare group AD severity categories according to the CDR so as to avoid arbitrary distinctions.

ADT cells were exposed for 24 h to FBS-free medium supplemented with serum from AD and MCI patients at 10% v/v concentration as described for PE serum samples. Intriguingly, our results revealed a large amount of protein aggregates as demonstrated by robust ProteoStat signal in the cells treated with serum samples from AD patients (n = 10) but not age-matched control serum samples (n = 19) (Fig. 4a). MCI is a pre-AD condition and would be expected to show equal or less content of protein aggregates compared to those with AD dementia. Thus, it is important to assess whether the ADT-ProteoStat assay can be applied to sera from MCI patients to detect similar protein components and content of aggregates at an earlier stage than observed in the AD serum samples. Accordingly, we tested serum samples from MCI patients (n = 14) who were diagnosed clinically as described above for the purpose of detecting protein aggregates. As shown in Fig. 4b, significant levels of aggregated proteins could be detected in the ADTs treated with MCI serum relative to age-matched controls. ROC analysis to distinguish cases from controls and MCI from AD revealed significant AUC values of 0.832-1.00 (Fig. 4c). It is important to point out that while the sample size is small, because of strict definition of clinical conditions in the cases and age-matched controls, we were nonetheless able to establish this novel discriminative assay. Furthermore, for AD and MCI serum samples, the research team was blind to group assignment (patients vs controls) supporting our rigorous approach to study design. The samples were decoded after completion of detection assays.

**Figure 4.**
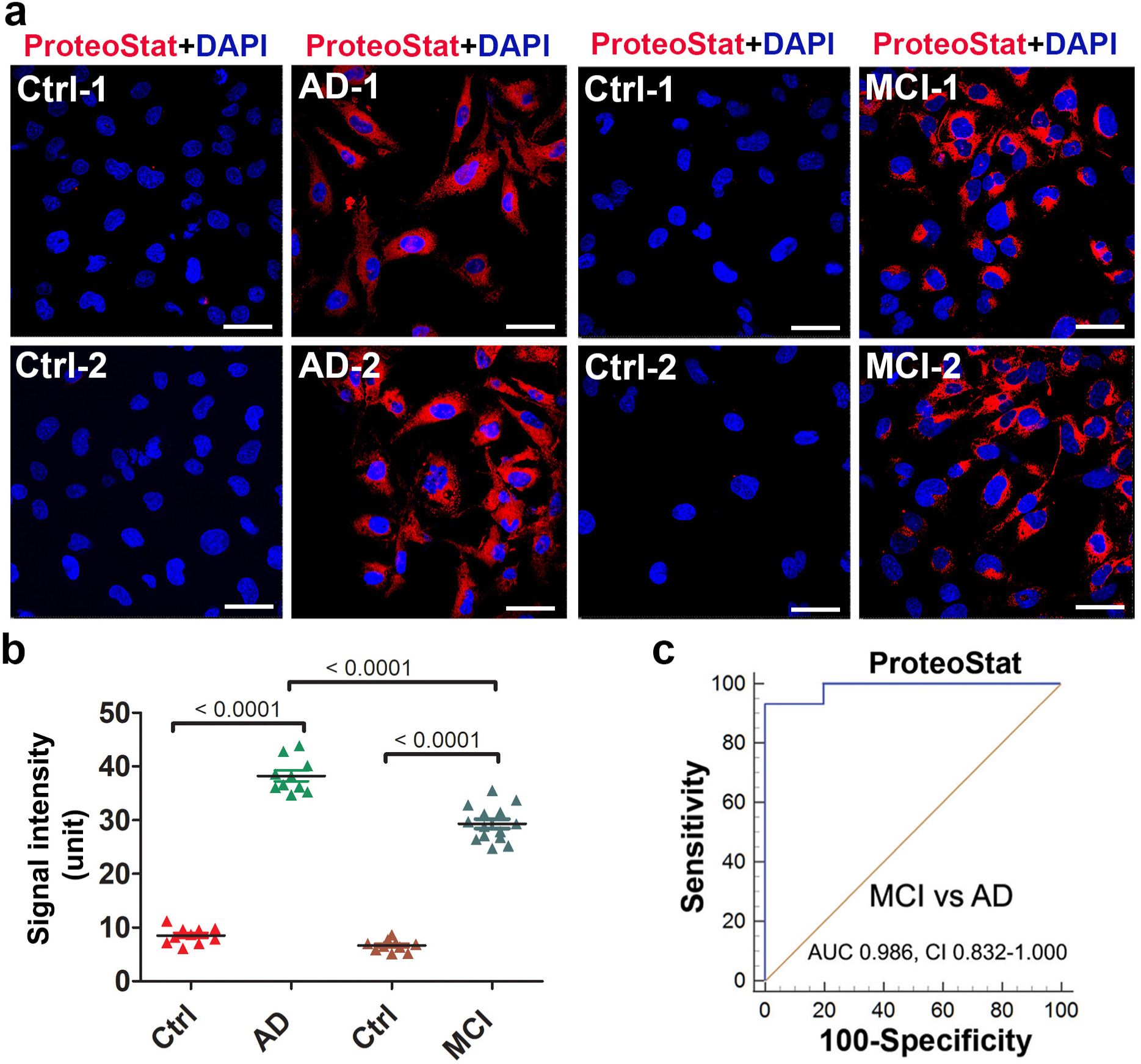
Detection of protein aggregates in sera from patients with mild cognitive impairment (MCI) and Alzheimer’s disease (AD). (A) ADTs were incubated with sera from MCI patients (n = 14), AD patients (n = 10) or their corresponding age-matched controls (Ctrl) (n = 10 for AD, n = 9 for MCI), fixed at 24 h and then stained with ProteoStat dye. The nuclei were stained with DAPI (blue). Bar: 50 µm. (B) The fluorescence intensity in cells were measured and statistically compared among groups. Data are expressed as mean ± SEM and analyzed by one-way ANOVA. (C) ROC analyses show robust difference in ProteoStat signal between the samples from MCI (n =14) and AD (n = 10) patients.

### Identification and validation of components of protein aggregates in sera from MCI and AD patients

To determine whether Aβ, phosphorylated tau protein, TTR, and α-syn are components of protein aggregates, we did a dual staining with specific antibodies and ProteoStat dye in MCI or AD serum-treated ADTs. Our results revealed a large amount of Aβ, TTR, α-syn (Fig. 5a and b) and co-localization of these proteins with robust ProteoStat signal in MCI or AD serum-treated ADTs, not with corresponding controls, demonstrating these proteins as the components of the aggregate complexes. Quantification analysis demonstrated that AD sera contained higher levels of Aβ and α-syn and a lower level of TTR aggregates than MCI sera (Fig. 5c-e). Fig. 5f shows the ROC curve analysis with robust differences in TTR, Aβ, and α-syn between MCI and AD serum samples. The AUC and CI values are all significant for TTR, Aβ and α-syn in distinguishing MCI from AD patients.

**Figure 5.**
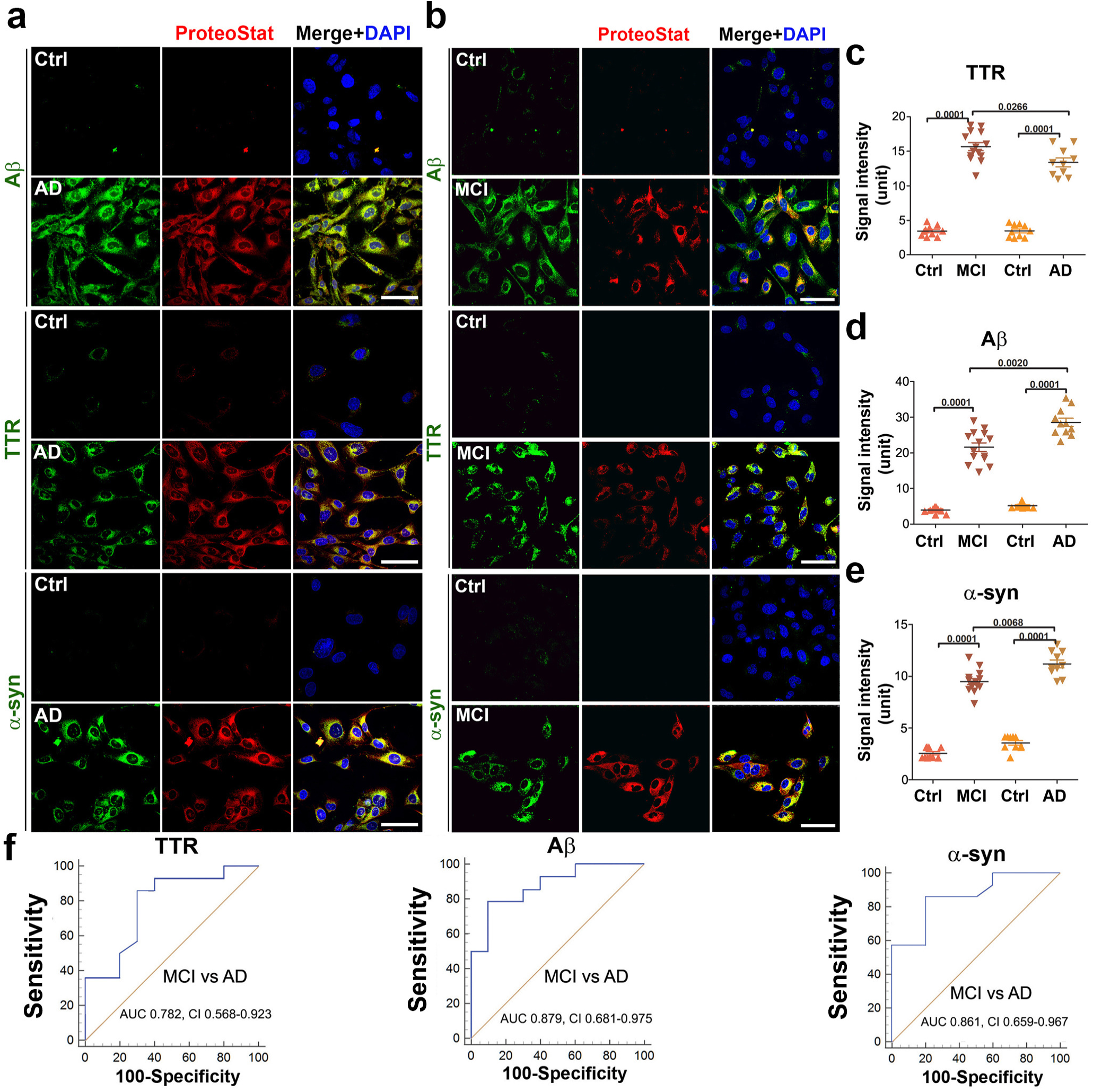
Identification of Aβ, TTR and α-synuclein as the components of the aggregates accumulated in ADTs treated with sera from AD or MCI. **(A, B)** Representative images show colocalization of TTR, Aβ or α-synuclein (α-syn) (green) with ProteoStat signal (red). ADTs were treated with 10% sera from AD (n = 10), MCI (n = 14) or control (n = 19), fixed at 24 h, immunostained with antibodies against Aβ, TTR or α-syn and then co-stained with ProteoStat dye. The nuclei were stained with DAPI (blue). Bar: 50 µm. (**C**) The fluorescence intensity of TTR, Aβ and α-syn in A and B was measured and statistically compared among groups. (**D**) ROC analyses show robust difference in TTR, Aβ and α-syn between the samples from MCI (n =14) and AD (n = 10) patients. Data are expressed as mean ± SEM and analyzed by one-way ANOVA.

The protein Tau harbors multiple potential phosphorylation sites, with the site T231 crucially important for the role of tau in regulation of microtubule binding and involvement in neurodegenerative diseases^52^. We are currently performing detailed analysis of different phosphorylated tau proteins in AD and MCI serum samples for quantitative differences and diagnostic value. For screening for tau proteins in this study, we focused on phosphorylated tau231 (T231). We observed robust phosphorylated tau (T231) immunofluorescence signal in ADTs exposed to sera from AD patients and only a moderate level of tau signal in MCI serum-treated cells (Fig. 6). Future experiments are currently being performed to characterize all potential tau proteins in AD and MCI serum samples.

**Figure 6.**
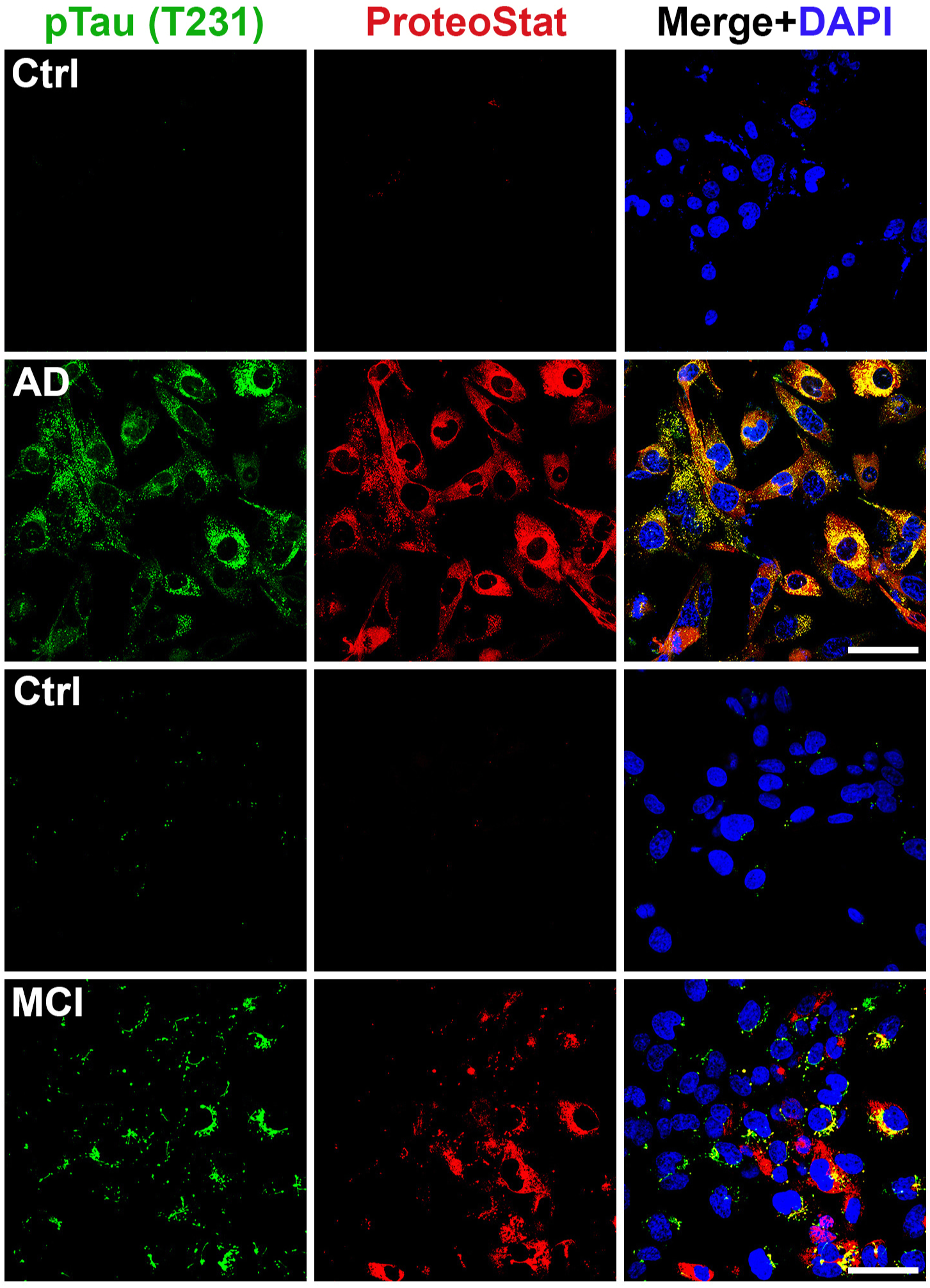
Detection of phosphorylated Tau231 (T231) in sera from patients with AD and MCI. ADT were incubated for 24 h with 10% of AD (n = 4), MCI (n = 4) sera or control sera (n = 4) and immunostained with specific anti-phosphorylated Tau (T231) antibody and co-stained with ProteoStat dye. Images are representatives of at least 3 independent experiments. Bar: 50 µm.

### Detection of serum-associated protein aggregates in ADT cells is not associated with transcriptional induction of protein components

Robust detection of protein aggregates in serum-exposed ADT cells raises the question whether mRNA for the individual protein components was induced in response to serum from PE or AD patients, which then contributed to excess protein production, leading to protein aggregation. We chose amyloid precursor protein (APP), TTR, and α-syn transcripts for analysis using real-time PCR (RT-PCR). Total RNA was extracted from ADTs treated with serum samples from AD and e-PE patients at 3, 12, and 24 hr. As shown in Fig. 7, exposure to PE or AD sera did not significantly alter the mRNA levels of these proteins over the time course (*p* > 0.05). This suggests that aggregated proteins are internalized from sera and accumulated over time inside ADTs rather than produced endogenously by the cells.

**Figure 7.**
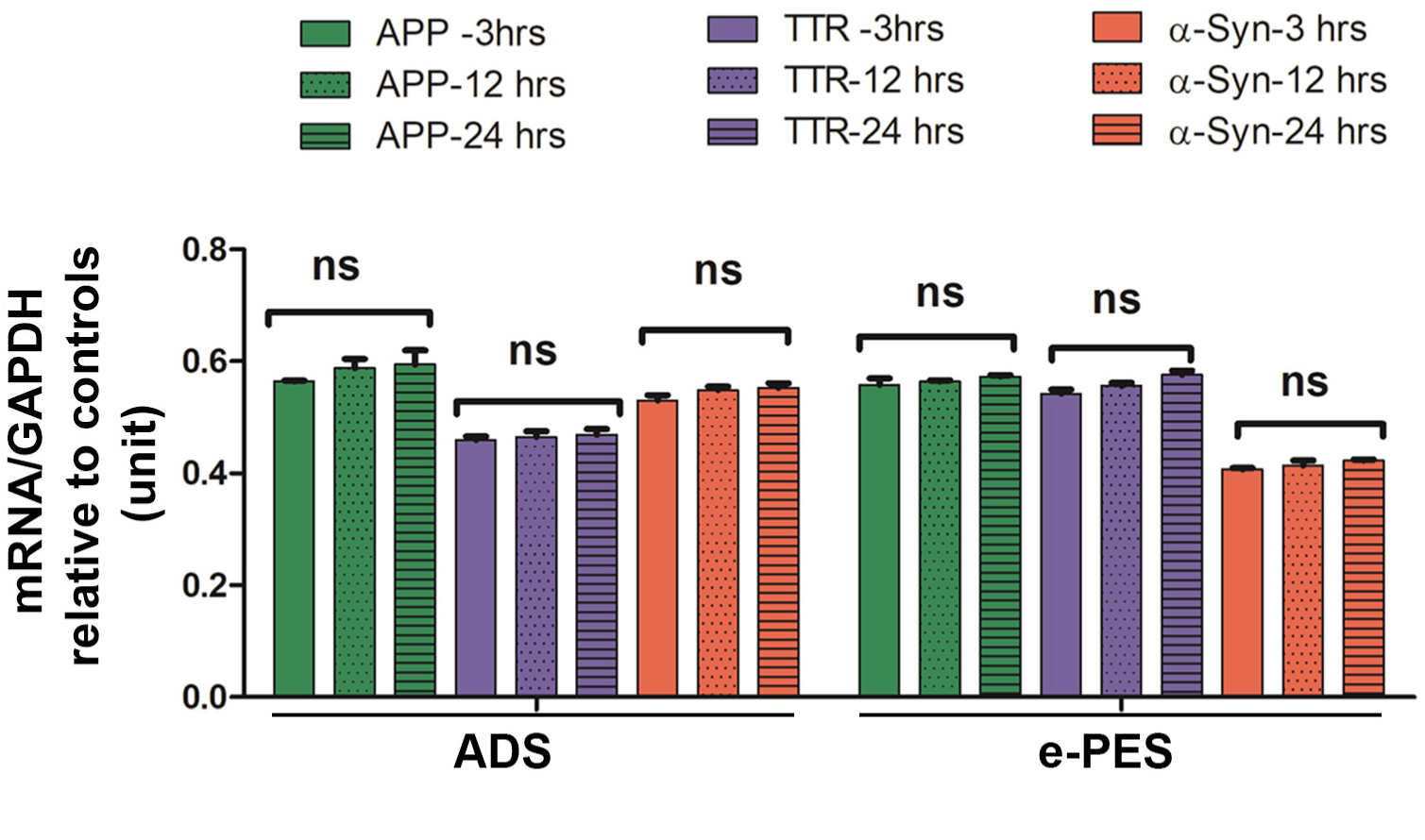
Time-course study of mRNA levels of indicated proteins in ADTs exposed to sera from AD or e-PE patients. ADT were incubated with ADS or e-PES, and total RNA was isolated at indicated time points and then RT-PCR was performed. The mRNA levels were normalized to house-keeping gene, GAPDH. The ratio of the mRNA levels in ADS- or e-PES-treated cells to those in respective control serum-treated cells was calculated to reflect the transcriptional alteration of each protein at each time point and then statistically compared among the groups. Data are expressed as mean ± SEM and analyzed by one-way ANOVA (n = 12). ns: not statistically significant.

## Discussion

Disease progression from preclinical to prodromal and dementia stages in AD is associated with accumulation of misfolded and aggregated proteins. In particular, in the case of AD and MCI, recent studies have focused on detection of individual blood biomarkers, including Aβ and phosphorylated tau proteins (pTau181 and pTau217), and on correlation of these findings with elevated levels of these proteins in CSF and increased tau PET and amyloid PET uptake^24, 38–42, 53, 54^. These observations are important in that they can correlate specific increases in phosphorylated tau proteins with the AD continuum. Similarly, the plasma Aβ42/Aβ40 ratio is considered to be an important marker for longitudinal progression to AD dementia^55^. Recently, α-syn has also been shown to be part of the AD pathology^29, 30^. While surpassing previous efforts to develop a blood test for AD, these newer proteomic tests do not detect aggregated proteins as part of a composite structure of biomarkers. To date, a rapid, sensitive, high through-put and cost-effective assay that can directly detect protein aggregates in fluids has been lacking.

We and others have shown that PE is another proteinopathy disease^5–11^. Several aggregated proteins have been detected in urine from women with severe PE using dot-blotting in combination with Congo Red staining^6^. However, since acute kidney injury usually occurs in PE patients, it is not clear whether protein aggregates come directly from the injured kidneys or from the placenta through circulation. During normal pregnancy, there is dynamic active transport of nutrients and protein molecules through the placental barrier at the maternal and fetal interface^46, 47^. Trophoblasts, as the frontline of the placental barrier that directly contact maternal blood, have the ability to internalize nutrients and large-sized protein molecules^46, 47^. Autophagy-lysosomal machinery is an essential biological process that degrades bulk proteins, such as misfolded protein aggregates or damaged organelles, to maintain cellular homeostasis^10, 11, 45^. Our prior studies have shown that impairment of the autophagy-lysosomal pathway is associated with the accumulation of protein aggregates in the trophoblast layer of the PE placenta^11^. Since ADTs exhibit impaired autophagy-lysosomal machinery^11, 45^, it was hypothesized that induced protein aggregates are not easily degraded and thus accumulate in a kinetic manner. Based on these observations, we developed a novel strategy for detecting protein aggregates in sera with the aid of engineered ADTs (Fig. 1). Our overall results confirm the validity of the assay and support the use of ProteoStat dye with its ability to bind to aggregated proteins. We have successfully applied this ADT-based protein aggregation assay to detect blood-based protein aggregates. Using this assay, we were able to detect aggregated proteins in sera not only from patients with e-PES and l-PES, but also now from AD and MCI in a high-throughput fashion.

Blood represents the most convenient fluid for a routine, non-invasive biochemical diagnosis for many diseases. On the other hand, blood is also a very complicated fluid containing high concentrations of albumin and immunoglobulin. These plasma proteins can easily form dense structures and may mask or interfere with protein aggregates at extremely low concentration in serum. Therefore, a challenging issue for detection serum protein aggregates is the requirement of high sensitivity for detection. Previous blood-based approaches need a high volume of plasma samples (e.g., up to 4 ml of plasma) to enrich or amplify target proteins through multiple complicated steps and expensive apparatus, and only detect single target protein^53^. In contrast, our assay is based on detection of total protein aggregates at a cellular level using ADTs, thus only requires a very small volume of serum. In our study, ADTs were cultured in medium with 10% serum in standard 12-well plates, and thus only 50 μl of serum was used in each sample assay. This can be further modified depending on availability of serum volume. Therefore, our ADT-based protein aggregation assay is highly sensitive. Moreover, our approach is cost effective as expensive devices are not required with the exception of a routine cell culture system and confocal fluorescence microscopy.

Notably, our assay, when combined with immunofluorescence staining, can be used to easily identify multiple specific proteins present in aggregates in one assay. By utilizing this method for the first time, we identified TTR and Aβ but not SERPINA1 and α-syn as major components of the aggregates observed in PE serum samples. These findings suggest that TTR and Aβ aggregates are released into maternal circulation from the PE placenta.

Importantly, our novel ADT-ProteoStat assay can be readily adapted as a potential predictive clinical assay using serum samples from preclinical stages for the onset of proteinopathy diseases in future studies. Early prediction of the onset of AD is crucially important for timely prevention and early intervention in this disease. However, the current tools that are used for identifying AD pathology pre-mortem, including brain tau and Aβ PET imaging and CSF protein analysis, are expensive, time-consuming and invasive^24, 38–42, 53, 54^. Although these methods have been used to predict pathology before clinical symptoms have developed, their invasiveness and cost have limited their use to well funded research settings in academic centers.

Using our novel assay, we identified TTR, Aβ, tau231 and α-syn as constituents of the aggregates seen in ADTs exposed to sera from patients with AD and MCI. Prior studies have shown that the concentration of Aβ and tau proteins is increased in the CSF or blood in AD^24, 38–42, 53, 54^. However, it was not clear whether these proteins are present as aggregates in biofluids. Using our assay, we were able to verify these proteins in the form of aggregates in the sera from AD and MCI.

While TTR aggregates are associated with multiple amyloid diseases including amyloidotic polyneuropathy, cardiomyopathy and systemic amyloidosis^8^, Native TTR itself may also serve as a neuroprotective molecule against AD^56–59^. Available evidence has shown that TTR binds to Aβ, prevents Aβ aggregation and clears Aβ through its proteolytic activity^56–59^. In support of this, a reduction of TTR content has been observed in CSF of AD patients^32^. Intriguingly, our assay revealed for the first time that TTR may exist as part of an aggregate complex together with tau, Aβ and a-syn in sera from MCI and AD. MCI is a transient condition that may precede probable AD later on^60^. To date, no biochemical test has been available for diagnosis of MCI. Our data suggest that aggregated serum proteins may be used as biomarkers for MCI. Therefore, our assay can potentially be used to predict the AD continuum.

The question arises as to whether the protein aggregates that accumulate in the ADT are internalized directly from sera or induced by pathological factors present in sera. Our observations indicate that the accumulated protein aggregates in ADTs are mainly derived from serum. First, we showed that ADTs had the ability to internalize and accumulate *in vitro* generated TTR aggregates. Second, depletion of aggregates from serum remarkably attenuated the accumulation of protein aggregates in ADTs and accumulation of aggregates occurred in a time-dependent fashion. Finally, no significant transcriptional alteration of TTR, α-syn and APP was observed in PE or AD serum-exposed ADTs, and α-syn only accumulated in ADTs when exposed to serum from AD and MCI patients not PE women.

Collectively, the development of a sensitive, reliable and generic protein aggregate detection assay for the early diagnosis of proteinopathy diseases is highly challenging. The results of this study support a novel, simple, and highly sensitive biomarker test that would be cost-effective in detecting total protein aggregates and individual proteins in serum at a cellular level. Given the small sample size in this study, future analysis of protein aggregates and their individual components in larger cohorts is warranted for better generalization and assessment of the sensitivity of this assay. To provide even more distinction between early and late onset PE or MCI and AD, a careful screening of serum samples of respective disorders at earlier time pints during incubation of ADT cells with serum is recommended which is likely to provide even more accurate information for clinical evaluation. Nonetheless, this new diagnostic technique is likely to be of great value for developing a point of care assay for detection of serum-based protein aggregates for PE and AD and can possibly applied to other proteinopathy diseases.

## Methods

### Human subjects for PE

This study was approved by the Institutional Review Boards at Women and Infants Hospital, Providence, RI and by the Regional Ethics Review Board, Linkoping, Sweden. Patients with PE were diagnosed based on ACOG guidelines of early onset and late onset preeclampsia. Systolic blood pressures ≥ 140 mmHg or ≥ 160 mmHg and diastolic blood pressure ≥ 90 mmHg or ≥ 110 mmHg measured at or after 20 weeks of gestation were associated with PE or severe PE, respectively. Serum samples were obtained following informed written consent from pregnant women with early onset PE (e-PE) (<34 gestational weeks), late onset PE (l-PE) (>34 gestational weeks), and gestational age-matched normal pregnancy. Exclusion criteria included chronic hypertension, gestational or pre-existing diabetes, fetal demise, daily tobacco use, fetal anomalies, and multiple gestations. For each study participant, 7-9 ml of blood was collected in BD Vacutainer SST™ tubes and processed for serum isolation within 30 minutes. Serum samples were aliquoted in smaller volumes and stored at -80 °C until further use. All methods were carried out in accordance with relevant guidelines and regulations.

### Human subjects for MCI and AD

This study was approved by the Lifespan Hospitals Institutional Review Board, Providence, RI. The Rhode Island Hospital Alzheimer’s Disease and Memory Disorders Center (ADMDC) Biospecimen Bank supplied the serum samples for the experiments. Upon enrollment, all participants give written consent for storage of biofluids for future research.

For this study, serum samples from patients with probable AD or MCI were age- and sex-matched to serum from normal controls (Supplementary Table 2). Patients were evaluated at the ADMDC clinic between 2010 and 2020 and had been diagnosed with AD based on National Institute on Aging-Alzheimer’s Association research criteria^43^, or with MCI, based on research diagnostic criteria^44^. Controls were healthy older adults, without cognitive impairment, recruited from among the clinic patients’ friends and family members. Blood samples were collected and processed according to the Alzheimer’s Disease Neuroimaging Initiative methods^44^, then plasma and serum components were aliquoted into 1 mL sterile polypropylene screw capped tubes and stored frozen at -80°C. Serum samples were de-identified prior to transfer and the investigators involved in running the experiments were blind to diagnostic group until completion of lab analyses.

### Autophagy-deficient and -proficient trophoblast cell line

The HchEpC1b cell line, HPV E6 and hTERT-transfected immortalized extravillous trophoblast cells, were stably transfected with pMRX-IRES-puro-mStrawberry-Atg4B^C74A^, an Atg4B^C74A^ mutant expression vector that inhibits MAP1LC3B-II formation, as previously described^45^. For the control, HchEpC1b cells were stably transfected with pMRX-IRES-puro-mStrawberry, a control vector only encoding monomeric red fluorescent protein^45^. After transfection, the cells were grown in RPMI1640 medium (GIBCO, 11875, MA, USA) supplemented with 10% FBS and selected by addition of 0.3 µg/ml puromycin (Sigma, P8833) in the medium. These cell lines were established by Dr. Akitoshi Nakashima (a co-author) and collaborators^45^.

### *In vitro* generation of TTR aggregates

Native human TTR solution (1mg/ml, AbD, Serotec) was diluted in sodium acetate buffer at pH 3.5 containing 100 mM KCl, 1 mM EDTA and 200 mM sodium acetate. We used a commercial kit to qualitatively measure protein aggregation of native and aggregated lysozyme as negative and positive controls along with BSA in this analysis. For a negative control, the same concentration of BSA solution was made with the same buffer. Both solutions were incubated in sealed tubes for 5-7 days at 37 °C. In pilot experiments, protein aggregates were qualitatively verified using a Protein aggregation assay kit according to manufacturer’s instruction (Enzo Life Sciences). Briefly, samples were mixed with ProteoStat dye and incubated for 10-15 mins at room temperature. Fluorescence signal intensity was read with a fluorescence microplate reader (Spectra Amax GEMINIEM, Molecular Devices) using an excitation setting of about 550 nm and an emission filter of about 600 nm.

### ADT-based protein aggregate detection assay

ADT or APT were plated on sterile glass coverslips and grown in RPMI1640 medium (GIBCO, 11875, MA, USA) supplemented with 10% FBS. After 65-70% confluency, the cells were washed with DPBS and then incubated in FBS-free RPMI1640 medium supplemented with 10% sera from women with e-PES, l-PES or their respective gestational age-matched controls, or sera from patients with AD, MCI or their respective age-matched controls. Treated cells were fixed at various time points with 4% formaldehyde in phosphate buffered saline (PBS) for 30 min at room temperature and then quenched with glycine for 5 mins. The cells were permeabilized for 30 min with a solution containing 0.5% Triton X-100 and 3 mM EDTA in PBS on ice. The cells were washed with PBS and then incubated with ProteoStat dye for 20 min at room temperature in darkness according to the manufacturer’s instructions (Enzo Life Sciences). The cells were then washed with PBS and mounted on glass slides with anti-quench mounting medium with DAPI (Vector Laboratories, Inc., Burlingame, CA) and observed using a confocal microscope (Nikon A1R, Japan) equipped with a 598 Red filter set. All images were acquired with a 60X objective lens. The signal intensity was measured using ImageJ software (NIH). Figures were processed with brightness/contrast adjustment using Photoshop CS2 (Adobe) using the same settings.

### Identification of the components of protein aggregates

The cells were treated for 24 h, fixed and permeabilized in a similar manner as described above. The cells were blocked by using a solution containing 1% BSA, 10% normal donkey serum and 0.3 M glycine in PBS for 1 hour at room temperature followed by overnight incubation with primary antibodies against TTR, Aβ, α-syn and SERPINA1 (1∶250 dilutions in blocking solution) in a humid chamber at 4 °C. The cells were washed three times in PBS buffer (pH 7.4) containing 0.1% Tween 20 (PBST) and incubated with Alexa Fluor 488 secondary antibody at 1∶500 dilutions in PBS containing 1% BSA for 2 h in dark. The cells were then counter-stained with ProteoStat dye (prepared according to kit instructions) for 30 min at room temperature. Finally, the slides were washed three times with PBST, mounted and observed under a confocal microscope (Nikon, A1R, Japan) using a Texas Red filter set for the ProteoStat dye and an FITC filter set for Alexa Fluor 488 conjugated antibodies. All images were acquired with a 60X objective lens. Negative controls were performed by replacing the primary antibody with purified rabbit IgG or mouse IgG. Immunoreactive intensity was measured with ImageJ (NIH). Figures were processed with brightness/contrast adjustment using Photoshop CS2 (Adobe) using the same settings.

### Immunoblotting

To discern the conformation of protein aggregation, equal amounts of protein extracts were separated with 4-15% SDS-PAGE under native conditions. Protein extracts were mixed with the sample buffer that does not contain reducing agents and SDS; the samples were not heated and were run in SDS-free Tris Buffer (Bio-Rad). After blocking in 5% nonfat dry milk dissolved in PBST for 1 h, the transferred PVDF membrane (Bio-Rad) was incubated overnight in primary antibody solution diluted in 5% nonfat milk or 3% BSA in PBST at 4 °C. After sufficient washes, the membrane was incubated for 1 h at room temperature with HRP-conjugated donkey anti-rabbit or mouse IgG (Cell signaling), treated with chemiluminescence substrate (SuperSignal, Pierce) and exposed on film (Kodak) or imaged using ChemiDo XRS^+^ (BIO-RAD). Density of bands was measured using ImageJ (NIH).

### Statistical analysis

Data were presented as the mean ± SEM and comparisons between experimental groups were statistically analyzed using a Student *t* test or one-way ANOVA followed by a post hoc test if *p* value is significant (GraphPad Prism Software, Inc). Differences between the groups were considered significant when the *p* value was < 0.05. Diagnostic accuracies were assessed with receiver operating characteristic (ROC) curve analysis using MedCalc Software version 19.3. The output included area under the curve (AUC), 95% confidence interval (CI) for the AUC, sensitivity, specificity, and significance level (*p* value).

### Data availability

The datasets generated and/or analyzed during the current study are available from the corresponding authors on reasonable request.

## Acknowledgments

We thank Paula Krueger for technical assistance. We also thank the Department of Pediatrics, Women & Infants’ Hospital of Rhode Island, Core Facility and Warren Alpert Medical School of Brown University, for continued support.

## Author Contributions

S.C., S.B., S.J., and Z.H. performed research. S.B., S.C., and S.S. (Surendra Sharma) designed research, performed data analysis and wrote the manuscript. A.N., S.S. (Shigeru Saito), J.P., B.O., L.D., J.D., J.E. and G.B. contributed reagents/samples/analytic tools and edited the manuscript. S.S. (Surendra Sharma) supervised the research.

## Funding

This work was supported in part by the NIH P20 GM121298, 3P20GM121298-04W1 and P30 GM114750 grants, Brown University DEANS Award, Brown University Seed Award, and William and Mary Oh-William and Elsa Zopfi Professorship Award.

## Competing interests

The authors declare no competing interests.

## Additional information

### Supplementary Information

The online version contains supplementary material available. Correspondence and requests for materials should be addressed to S.S.

---

**Figure S1.**
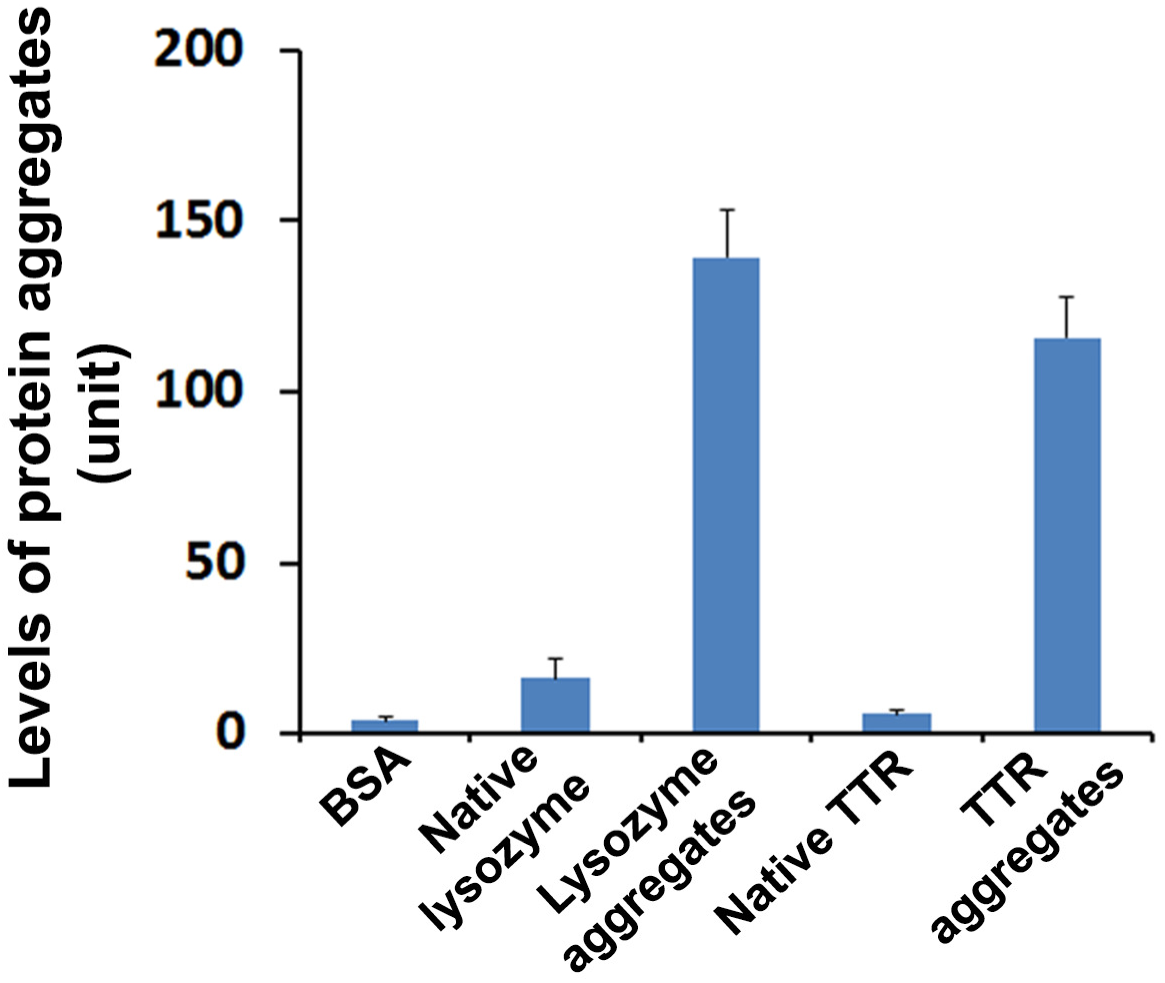
Validation of *in vitro* generated TTR aggregates. Strikingly higher levels of protein aggregates were detected in the solution containing our *in vitro* generated TTR aggregates and commercial lysozyme aggregates (Enzo) compared with BSA, native lysozyme and native TTR (*p* < 0.001). Protein aggregates were detected from indicated samples using ProteoStat protein aggregation assay kit.

**Figure S2.**
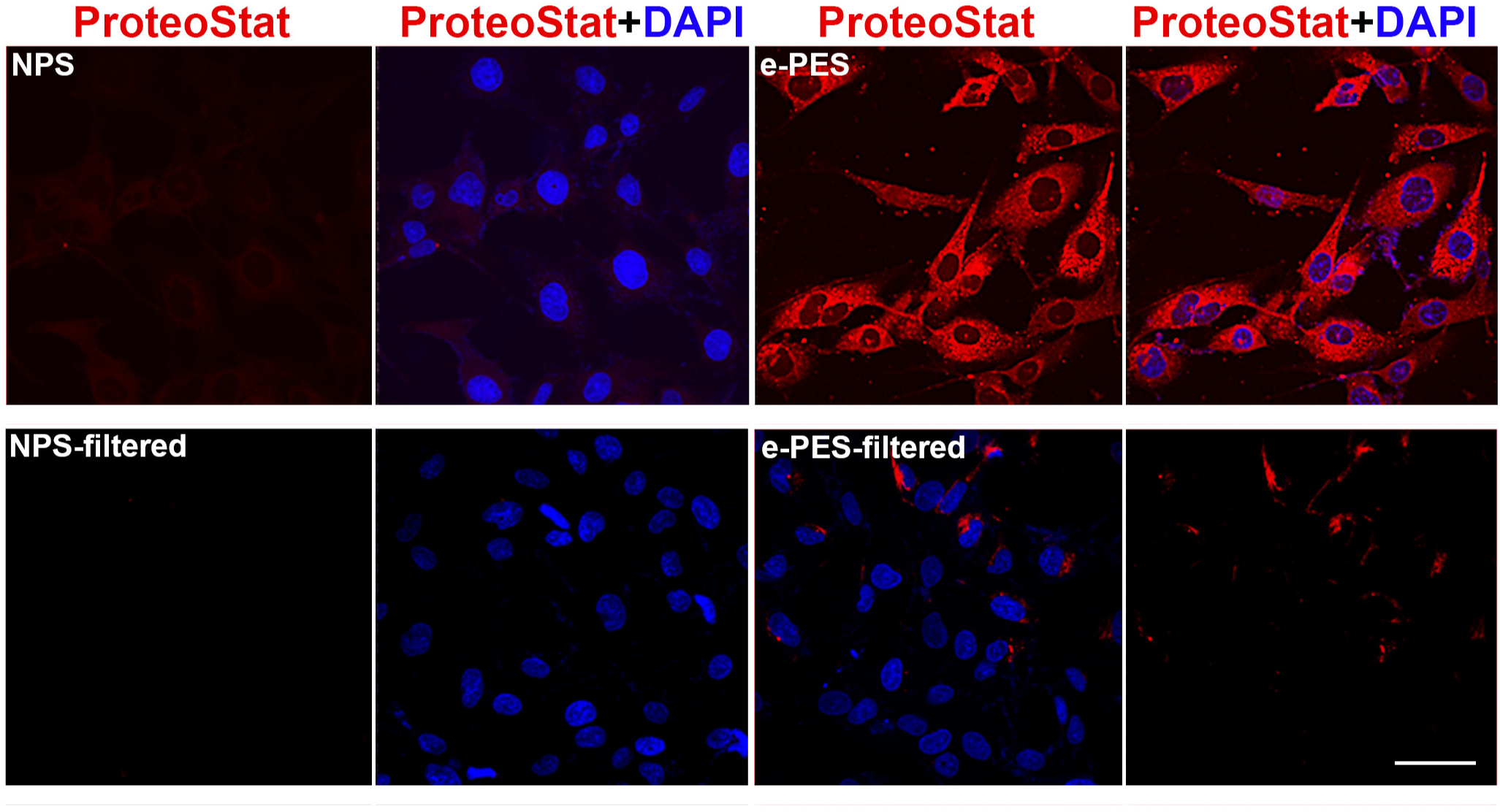
Depletion of the aggregates from e-PE sera attenuates the accumulation of protein aggregates in ADT. Cells were incubated for 24 h with e-PE sera or control sera before and after depletion of aggregates, fixed and then stained with ProteoStat dye. Images are representatives of at least 3 independent experiments. The nuclei were stained with DAPI (blue). Bar: 50 µm.

**Figure S3.**
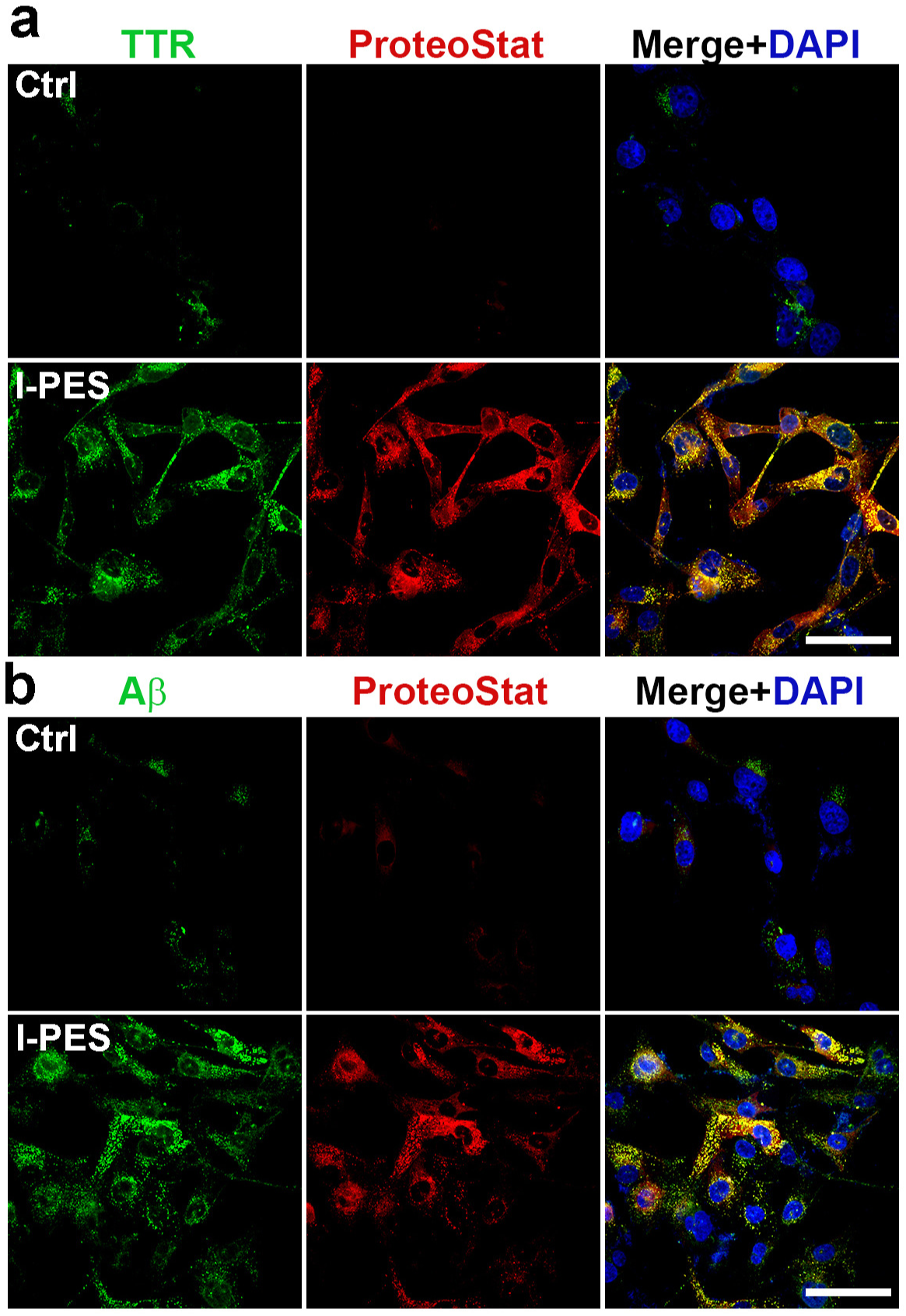
Identification of TTR and Aβ as components of the aggregates in ADT exposed to sera from l-PE (l-PES). ADT were incubated with l-PES or control sera (Ctrl), fixed at 24 h and immunostained for TTR (green, **A**) or Aβ (green, **B**) and then counter-stained with ProteoStat dye (red). The nuclei were stained with DAPI (blue). Images are representatives of at least 3 independent experiments. Bar: 50 µm.

**Figure S4.**
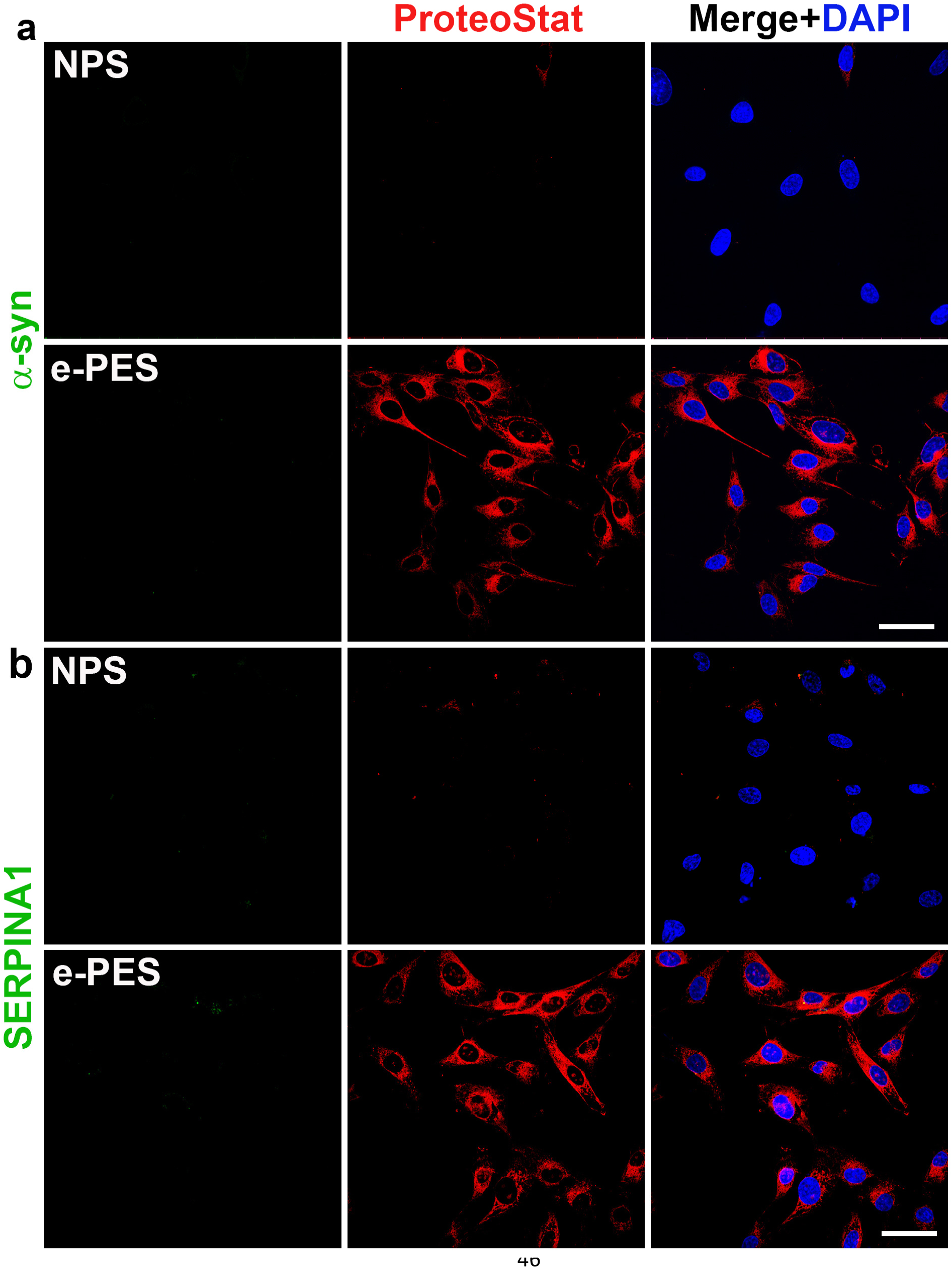
Aggregated α-synuclein and SERPINA1 are not detected in e-PE serum-treated ADT. ADT were incubated with e-PE sera (e-PES) or control sera (NPS), fixed at 24 h, immunostained for α-synuclein (α-syn, **A**) or SERPINA1 (green, **B**) and co-stained with ProteoStat dye (red). The nuclei were stained with DAPI (blue). Images are representatives of at least 3 independent experiments. Bar: 20 µm.

**Table S1.**
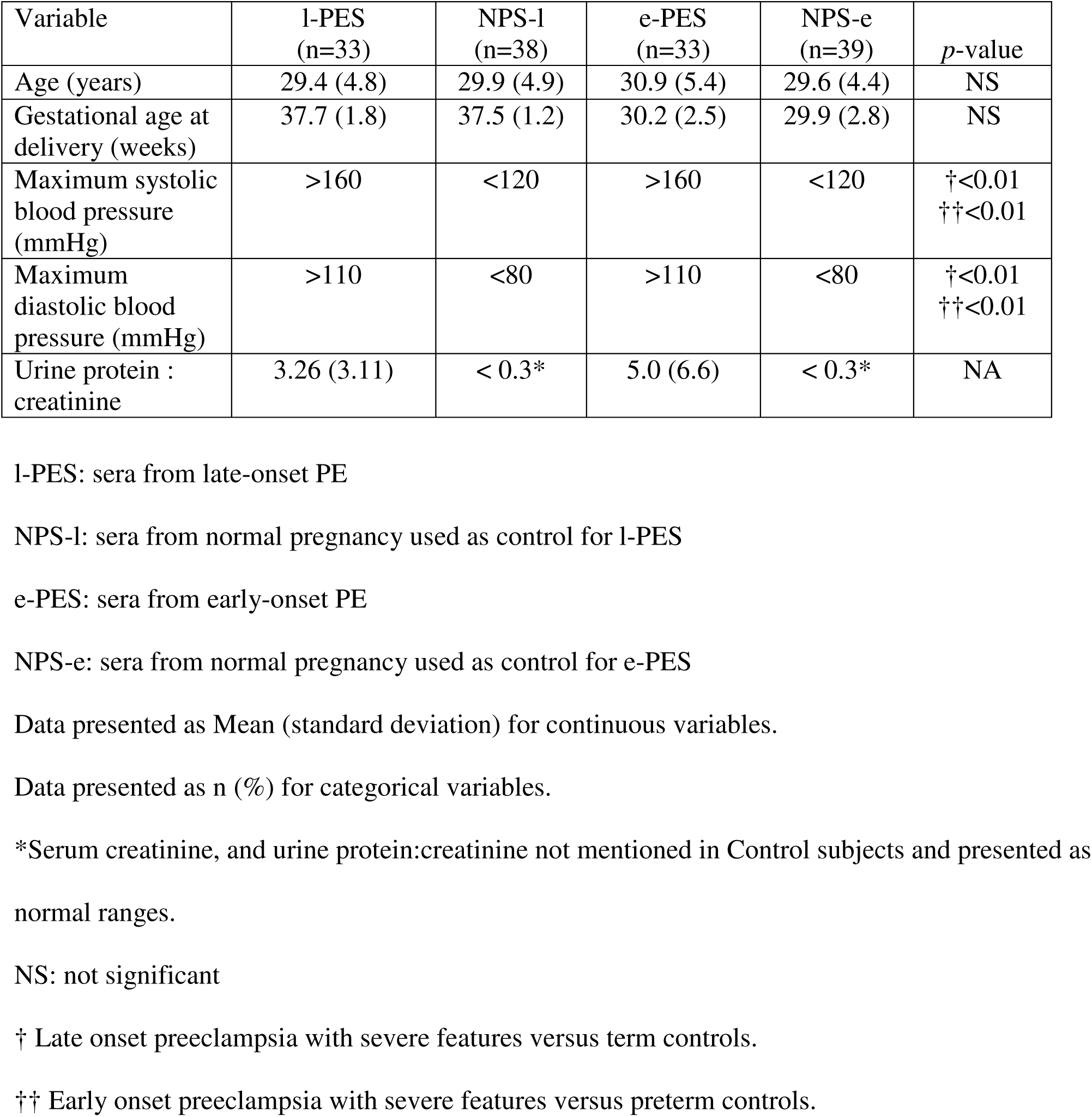
Demographic data of patients with early-/late-onset preeclampsia and control pregnancies

**Table S2.** Demographic data of patients with MCI and AD and age-matched cognitively normal controls

|  | Cognitive Normals<br>(Controls)<br>n = 19 | MCI<br>(Possible<br>AD)<br>n = 14 | AD Dementia<br>(Probable AD)<br>n = 10 |
| --- | --- | --- | --- |
| Age, mean, yrs (SD) | 70.6 (9.3) | 70.1 (8.5) | 69.8 (9.5) |
| Sex, F/M, n | 13/6 | 8/6 | 6/4 |
| Education, mean, yrs (SD) | NA<sup>†</sup> | 14.5 (2.8) | 14.3 (2.0) |
| MMSE, mean, score (SD) | NA | 26.5 (2.2) | 16.8 (7.4) |
| CDR, 0.5/1/>1, n | NA | 12/0/0 | 0/5/2 |
| AD Biomarkers*,<br>Amyloid PET+/CSF+/FDG-<br>PET +, n | NA | 11/1/0 | 3/2/1 |

AD: Alzheimer's Disease; SD: standard deviation; MMSE: Mini-Mental State Exam, range 0-30; CDR: Clinical Dementia Rating Scale, range (0-3); PET: Positron Emission Tomography; FDG-PET: <sup>18</sup>F-fluorodeoxyglucose positron emission tomography
\* AD Biomarker Positive (+) indicates: a) amyloid plaque burden (PET brain imaging), b) cerebrospinal fluid concentrations of amyloid-beta (Aβ <sub>1-42</sub>), total tau (T-tau), and phosphorylated tau (P-tau<sub>181</sub>) within established reference ranges for MCI or AD dementia, or c) neuronal dysfunction measured by FDG-PET. Number of subjects (n) who underwent biomarker analysis in each category is reported.
<sup>†</sup> Not assessed (NA)

## Notes

### Competing Interest Statement

The authors have declared no competing interest.

### Author Declarations

This study is case control study and was approved by IRB committee of Women and Infants Hospital of Rhode Island (Project ID# WIH-02-0061). Informed written consent was obtained from each subject.

